# Reparations for Black American Descendants of Persons Enslaved in the U.S. and Their Estimated Impact on SARS-CoV-2 Transmission

**DOI:** 10.1101/2020.06.04.20112011

**Authors:** Eugene T. Richardson, Momin M. Malik, William A. Darity, A. Kirsten Mullen, Maya Malik, Aletha Maybank, Mary T. Bassett, Paul E. Farmer, Lee Worden, James Holland Jones

## Abstract

**Background:** In the United States, Black Americans are suffering from significantly disproportionate incidence and mortality rates of COVID-19. The potential for racial-justice interventions, including reparations payments, to ameliorate these disparities has not been adequately explored.

**Methods:** We compared the COVID-19 time-varying *R_t_* curves of relatively disparate polities in terms of social equity (South Korea vs. Louisiana). Next, we considered a range of reproductive ratios to back-calculate the transmission rates *β_i_*_→_*_j_* for 4 cells of the simplified next-generation matrix (from which *R*_0_ is calculated for structured models) for the outbreak in Louisiana. Lastly, we modeled the effect that monetary payments as reparations for Black American descendants of persons enslaved in the U.S. would have had on pre-intervention *β_i_*_→_*_j_*.

**Results:** Once their respective epidemics begin to propagate, Louisiana displays *R_t_* values with an absolute difference of 1.3 to 2.5 compared to South Korea. It also takes Louisiana more than twice as long to bring *R_t_* below 1. We estimate that increased equity in transmission consistent with the benefits of a successful reparations program (reflected in the ratio *β_b→b_* / *β_w→w_)* could reduce *R*_0_ by 31 to 68%.

**Discussion:** While there are compelling moral and historical arguments for racial injustice interventions such as reparations, our study describes potential health benefits in the form of reduced SARS-CoV-2 transmission risk. As we demonstrate, a restitutive program targeted towards Black individuals would not only decrease COVID-19 risk for recipients of the wealth redistribution; the mitigating effects would be distributed across racial groups, benefitting the population at large.

**Funding:** ETR and LW are supported by NIGMS MIDAS grant R01 GM130900. ETR is also supported by NIAID K08 AI139361. WAD is supported by NIMHD R01 MD011606, NSF SES 1851845, and IES R305A190484. MMM is supported by the Ethics and Governance of Artificial Intelligence Fund.

## Background

The novel coronavirus which causes COVID-19 was first reported in Hubei Province, China in December 2019.^1^ In the ensuing 5 months, the outbreak spread to nearly every country in the world.^2^ As of 28 May 2020, the United States had the highest number of reported cases of COVID-19 with 1,698,523 confirmed infections and 100,446 total deaths^3^—although this certainly represents an underestimate of the true number of cases given the poor scale-up of testing coupled with a high rate of asymptomatic infection.^4,5^

As has been the case in previous pandemics, communities of color are suffering from disproportionate incidence and mortality rates of COVID-19.^6,7^ In the states that have released data by race, this gap is notably pronounced among Black Americans who are dying at an age-adjusted rate that is 3.6 times as high as the rate for whites.^8^ It further adds to vast disparities in Black and white health that have been the cumulative result of legacies of slavery, legal segregation, white terrorism (e.g., lynchings during the Jim Crow period), hyperincarceration, lethal policing, and ongoing discrimination in housing, employment, policing, credit markets, and health care.^9–11^ While frameworks for understanding the mechanisms through which biosocial forces become embodied as pathology are inchoate, allostatic load (the physiological profile influenced by repeated or chronic life stressors) can be used to demonstrate how the continuous trauma of oppression can lead to disparities in health by race.^12–19^ The potential for racial-justice interventions, including reparations payments, to ameliorate these disparities, has not been adequately explored.^20^

Mathematical and computational models of infectious disease transmission dynamics increasingly are being used to determine the potential impact of interventions on incidence and mortality.^21^ Fundamental to this work is calculation of the basic reproductive number *R*_0_, which is defined as the expected number of secondary cases caused by a typical infected individual in a fully susceptible population.^22^ While *R*_0_ provides theoretical information about an epidemic, practical control ultimately depends on the expected infections generated later in the outbreak prompting epidemiologists to utilize the effective reproduction number *R_t_* (i.e., the average number of secondary cases generated by an infectious individual at time *t*), which obviates the assumption of a fully susceptible population and allows for the temporal dynamics to be followed in the setting of various interventions.^23^

Models must make assumptions about how people interact with others,^24^ but they rarely account for social forces like institutional and cultural racism that structure such interactions.^25^ Therefore, they can obfuscate such forces in their attempts to describe outbreak transmission dynamics.^26–28^

Nonetheless, it is possible to incorporate risk heterogeneities into models, and to use this information to identify more just measures for disease prevention/control.^29–33^ For example, Black workers are overrepresented in front-line sectors like food service and delivery, healthcare, and child-care, which places them at higher risk of SARS-CoV-2 infection. Furthermore, Black individuals have a higher likelihood of living in dense, precarious housing where effective social distancing is hindered. These risks are structural—that is, not determined by personal choice or rational assessment, as models often assume (what Koopman and Longini refer to as “the erroneous attribution of individual effects”^34^);^35,36^ they could therefore benefit from structural interventions. As such, the following modeling study explores the potential effects of reparations payments on the disproportionate COVID-19 risk among Black people in the U.S.

## Methods

For a representative inegalitarian state in the U.S., we chose Louisiana as a unit of analysis due to the availability of COVID-19 data compiled by race. Louisiana has one of the highest GINI coefficients (a measure for household income distribution inequality—a value of 0 represents total income equality [all households have an equal share of income], while a value of 1 represents total income inequality [one household has all the income]) among the American states (0.5),^37^ is highly segregated between Black and non-Black populations (Supplemental Table 1), and has significant differences in the average number of persons per room (PPR) for Black and non-Black populations (Figure 1). (PPR is a measure of overcrowding^38^ that recent reports indicate might be a more important for risk of infection than urban density.^39^)

**Figure 1.**
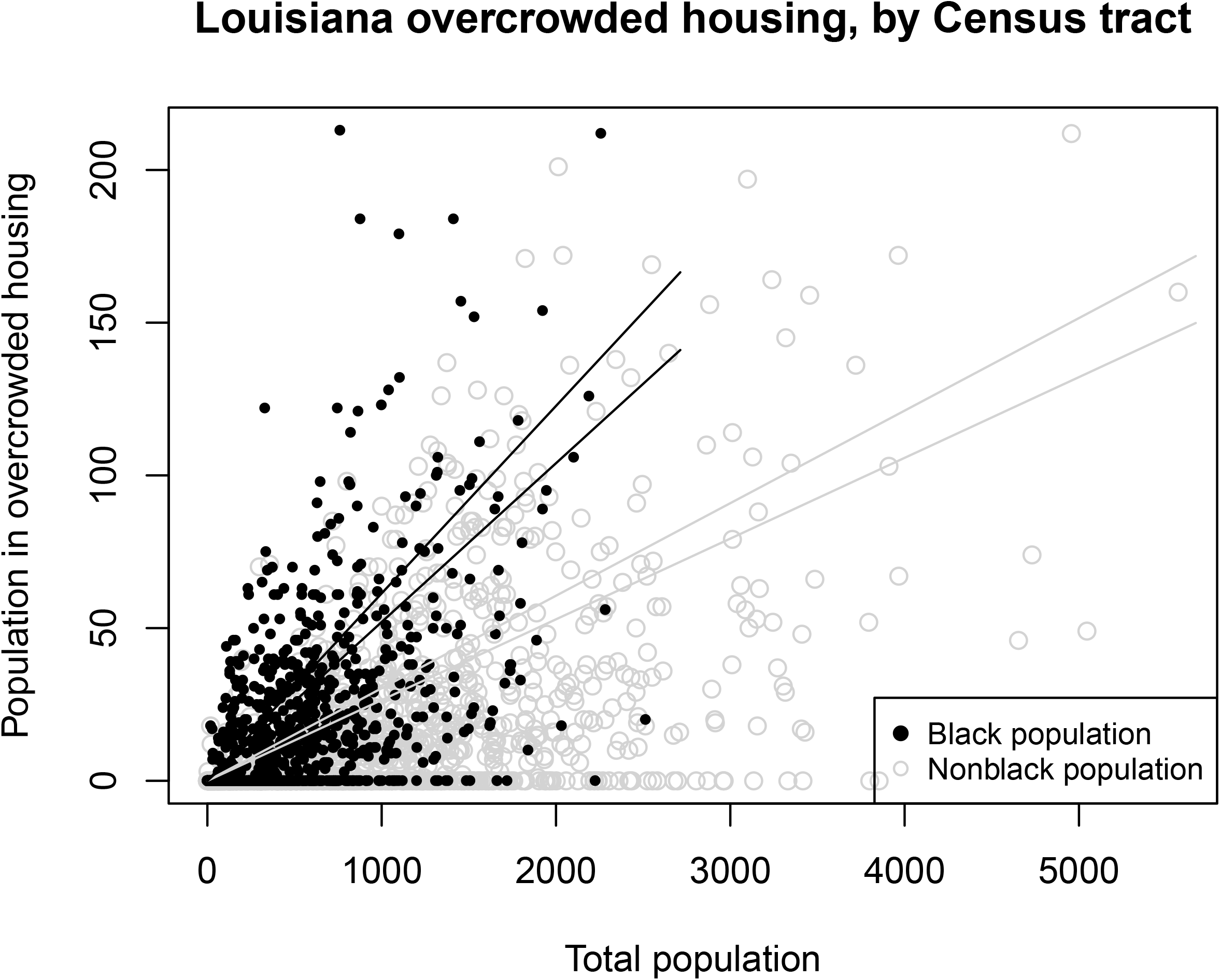
Fitted 95% confidence intervals for the ratio of the population in overcrowded housing to total population by Louisiana Census tract. In tracts where overcrowded housing is documented, the estimated ratio of Black population in overcrowded housing to total Black population is 0.0565, double that of the non-Black population (0.0283).

As of 28 May 2020, the state reported 38,802 SARS-CoV-2 infections.^40^ We estimated time-varying (instantaneous) *R_t_* using the method of Wallinga and Teunis,^41^ which uses a probability distribution for the serial interval (i.e., the time, in days, between symptom onset in an index case and symptom onset in a person infected by that index case). Confidence intervals for *R_t_* were calculated using a normal approximation for the estimated number of secondary cases per case (i.e. approximating the 95% central interval by the expected value *R_t_* +/- 1.96 times the standard deviation). Following current best estimates of the serial interval, we chose a gamma distribution with a mean of 5 and standard deviation of 2.^42,43^

To juxtapose these data with those reported from a relatively egalitarian polity, we conducted a similar analysis of the outbreak in South Korea,^44^ which in contrast to Louisiana, has a GINI coefficient of 0.32^45^ and no large, segregated subgroup of the population composed of the descendants of enslaved persons. South Korea nonetheless has 10 times the population density of Louisiana such that, if density *per se* were the major determinant of epidemic severity, we would expect rates of infection to be much higher in the former compared to the latter, which is not the case (Louisiana has reported nearly 40 times the number of cases per 100,000 people as South Korea).^40,44,46,47^

To estimate the effect prior reparations payments may have had on the outbreak in Louisiana, we considered reproductive ratios from the range of pre-intervention *R_t_* (a stay-at-home order was issued on 22 March 2020). From the theory of epidemics in structured populations, we know that *R*_0_ is given by the dominant eigenvalue of the next-generation matrix, **G**, a *k* x *k* matrix (Equation 1) that accounts for the movement into and between the *k* infection states in the population. The matrix is comprised of elements *g_i_*_→_*_j_*, which are the expected number of type-*j* cases caused by contact with infectious individuals of type *i*.^48^ With the assumed values of *R*_0_, we could analyze risk structure by back-calculating the transmission rates *β_i→j_* (which are equal to the contact rate *c_i→j_* multiplied by the transmissibility *τ* probability of infection given contact between a susceptible and infected individual]—Equation 2) for 4 cells of the next-generation matrix, with the following constraints: *b→b* >> *w→w* >> *w→b→* > *b→w* (where *b* = Black and *w* = white); the quantity *β_w→w_n_w_*/*γ*, which is the expected number of secondary cases caused by a typical infected individual within the white subgroup, was held constant at 1.5 (the median *R_t_* value for the first 2 weeks of the South Korea outbreak once daily reported cases were greater than 50^49^); the population proportion *π* was determined using data from the U.S. Census’ American Community Survey 5-year estimates for 2018 (Black = 0.36 and white/other = 0.64); and the recovery rate *γ* (the inverse of the infectious period) was held constant at a value of 0.06.^50^

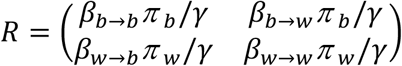

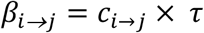

Lastly, we modeled the effect a program of reparations for Black American descendants of persons enslaved in the U.S. (i.e., total payments in the amount of $250,000 per individual or $800,000 per household^51^) would have had on pre-intervention *c_i→j_* and *τ* (see Supplementary Material Section 3 for more detailed methodology). We assume that post-reparations *R*_0_ for the population would have a lower bound of 1.5 (not a mean) since wealth redistribution would decrease the ability of affluent whites to cloister themselves in a setting of relative exclusivity.

We conducted all analyses in R (v. 4.0.0) for Macintosh, R Foundation for Statistical Computing, Vienna, Austria) using code written by the authors.

## Results

Figure 2 presents time-varying *R_t_* for the COVID-19 outbreaks in Louisiana and South Korea on a common scale of epidemic-days. Once the respective epidemics begin to propagate Louisiana displays *R_t_* values with an absolute difference of 1.3 to 2.5 compared to South Korea. It also takes Louisiana more than twice as long to bring *R_t_* below 1, the critical value at which an outbreak will die out in a population (24 days vs 11).

**Figure 2.**
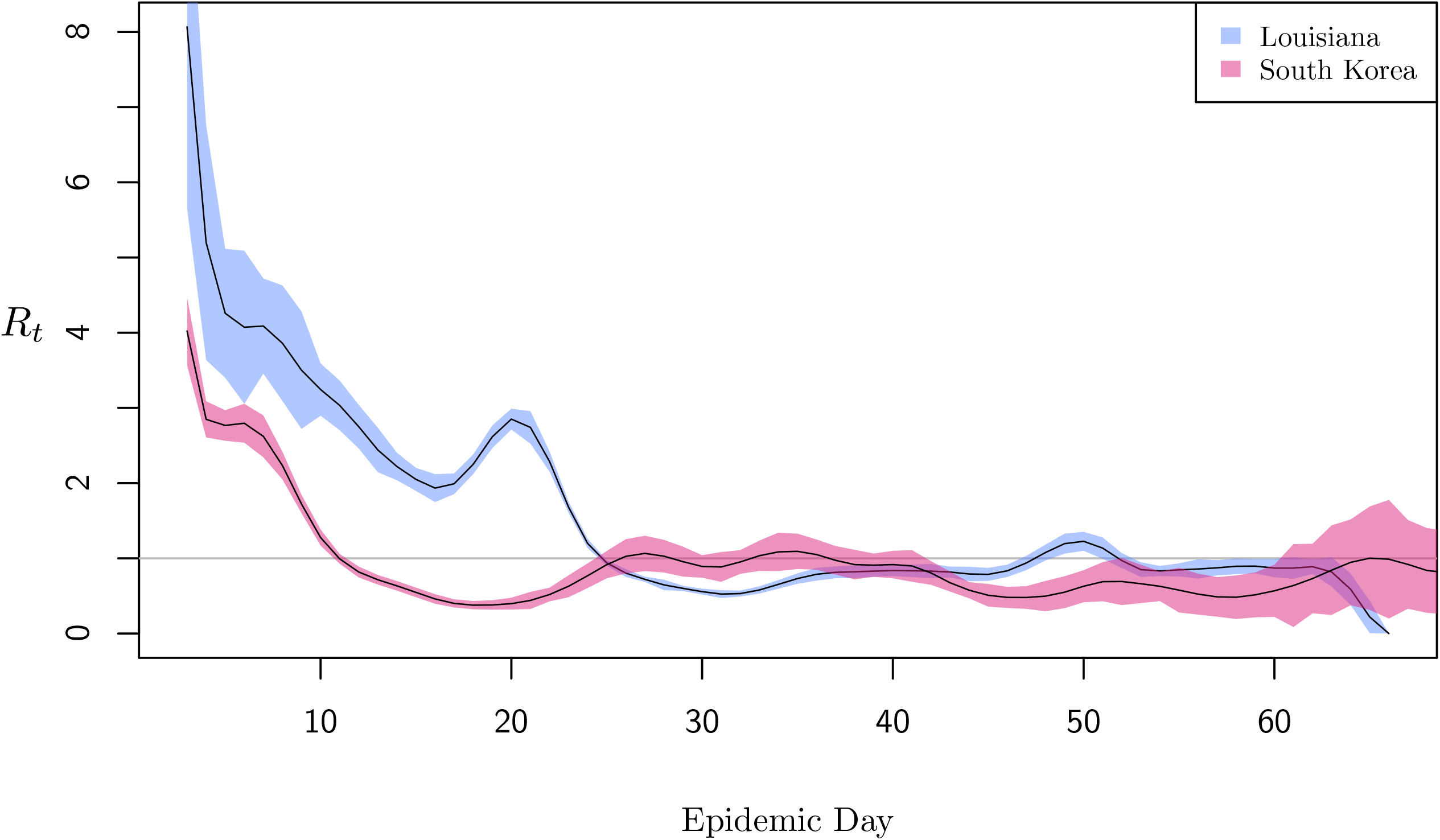
Estimated time-varying *R_t_* for the COVID-19 outbreaks in Louisiana (purple) and South Korea (red). Shaded areas represent 95% confidence intervals.

Our next-generation matrix analysis shows that, in a segregated society like the U.S. where SARS-CoV-2 transmission rates are disproportionate across racial groups, small changes in the ratio between *β_b→b_* and *β_w→w_* can result in large changes in the reproductive ratio for the population (Figure 3a), due mainly to 1) the effects of high assortative mixing structured by racism on the value of *c_b→b_*; and 2) the fact that the expected number of secondary infections generated within high-risk subgroups (i.e., the value *g_b→b_* in the next generation matrix—in this case driven by high relative values of *c_b→b_)* comes to dominate *R_0_* for a population.^29,52^

**Figure 3a.**
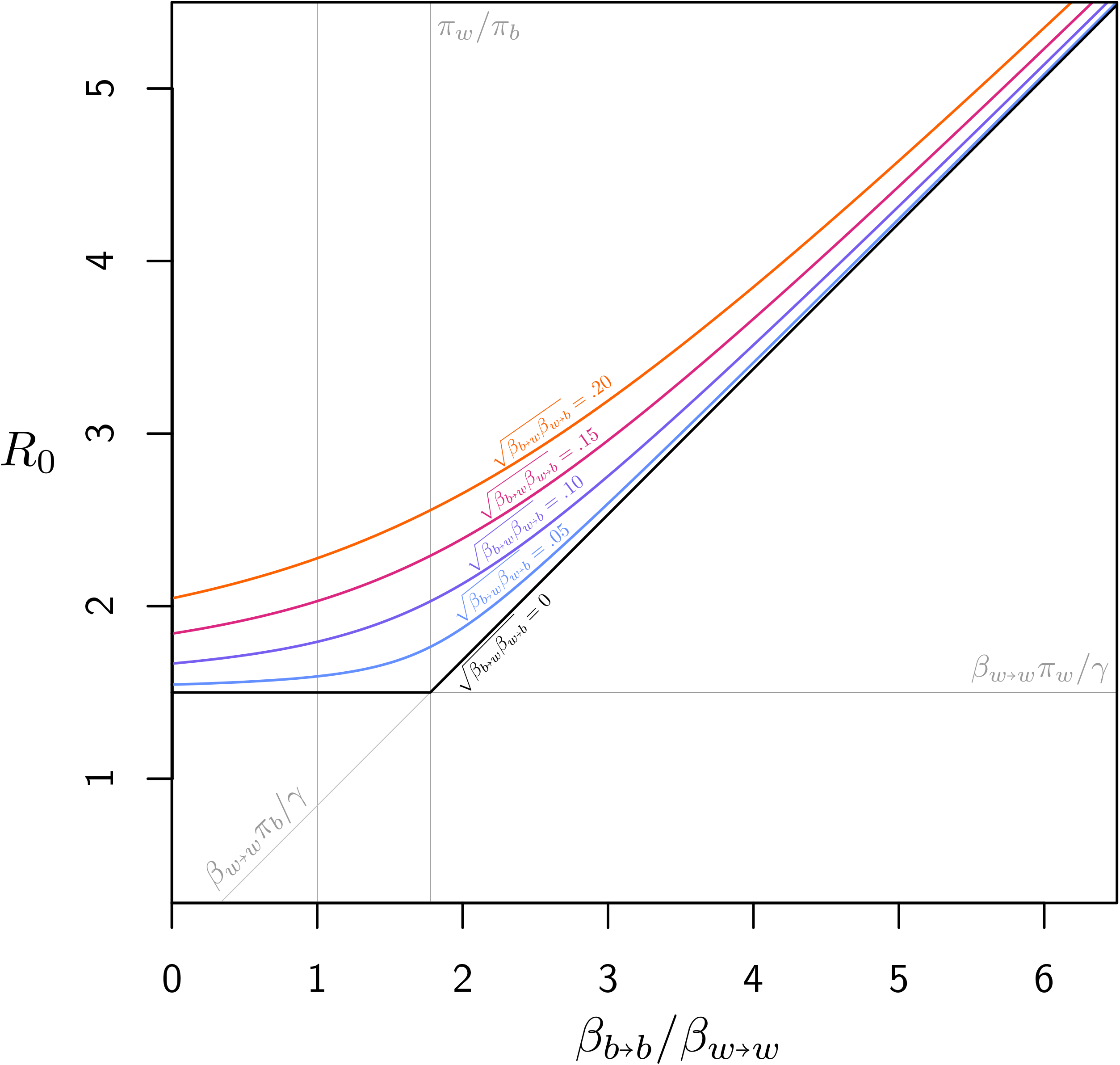
If we assume the lower bound on the basic reproduction number reflects the risk of the non-Black population, then to explain an estimated *R_t_* of 4, *β_b→b_* would have to be more than quadruple that of *β_w→w_*. This holds across different values of the off-diagonal terms (which affect the curve only through their geometric mean *(β_b→w_ β_w→b_)*^1/2^.

A program of reparations has the potential to reduce several variables that determine the COVID-19 reproductive ratio in such a segregated society. These include

i. reducing *c_i→j_* significantly for Black people by decreasing overcrowded housing (this also has the benefit of improving an individual’s ability to social distance once stay-at-home orders are enacted);
ii. reducing *β_w→b_* as Black individuals would not be forced as frequently into high-risk frontline work—with both attendant exposure and psychosocial stress;
iii. decreasing *τ* slightly on account of people’s ability to access preventive modalities like masks, hand sanitizer, etc.

Accordingly, the arrow in Figure 3b shows how different assumptions regarding the effects of reparations could play out: It begins within the range of *R_0_* we selected from the Louisiana outbreak pre-intervention (i.e., before the stay-at-home order was enacted); it ends within our estimates for *R_0_* in the setting of reparations, which are consistent with early values of *R_t_* estimated for South Korea and are 31 to 68% lower than pre-intervention estimates for Louisiana. This is achieved by the transmission rate *β_b→b_* decreasing to near parity with *β_w→w_*, which reflects the anticipated mitigation in structural racism a successful reparations program would engender.

**Figure 3b.**
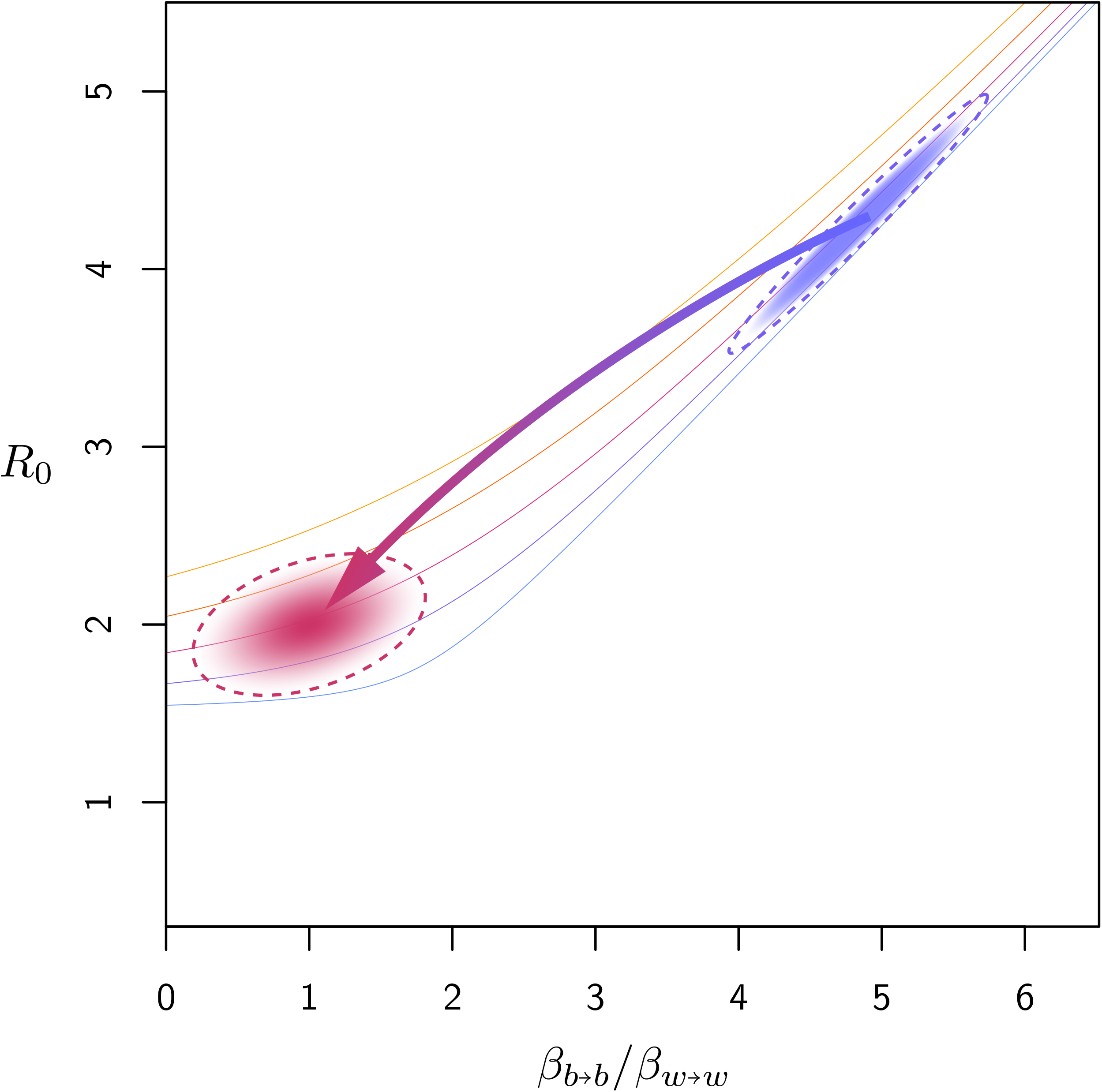
Arrow shows the potential effects of reparations for a range of *R_0_* derived from the Louisiana outbreak. Arrowhead ends in a shaded area that represents the range of transmission ratios *(β_b→b_* / *β_w→w_)* assumed to obtain from a successful reparations program, and the decreases in *R_0_* that would result.

## Discussion

### The Color Line

In the United States, where the problem of the 21st century is still the problem of the color line, 400 years of structural racism, violently-seized privilege, and continuous trauma from racial terror and dehumanization are clearly manifested in the disproportionate incidence and mortality rates of COVID-19 among Black Americans.^53,54^ While there are compelling moral and historical arguments for racial injustice interventions such as reparations,^55,56^ our study describes potential health benefits in the form of reduced SARS-CoV-2 transmission risk. As we demonstrate, a reparative program targeted towards Black individuals would not only decrease COVID-19 risk for recipients of the wealth redistribution; the mitigating effects would be distributed across racial groups, benefitting the population at large. Populations become entrained by the dynamics of the highest-risk segment of the population.^57^ This can be seen when a single diagonal element of the next generation matrix dominates the eigenstructure of that matrix. Overall, reducing the infectious burden on the most at-risk segment of the population has the greatest impact on the epidemic for the population at large. Reducing severe inequalities is thus not simply just, it is epidemiologically efficacious for outbreak containment.

In general, a program of reparations is intended to achieve three objectives: acknowledgment of a grievous injustice, redress for the injustice, and closure of the grievances held by the group subjected to the injustice.^58^ Potential mechanisms by which reparations—through both monetary compensation and acknowledgement of injustice—could have suppressed the world’s largest outbreak of SARS-CoV-2 include

1. narrowing of the path-dependent racial wealth divide;
2. changes in the built environment, fostering the ability to social distance;
3. spreading out of front-line work across racial groups;
4. decreased race-based allostatic load.

### Disproportionate Incidence vs. Mortality

Current explanations of excess COVID-19 risk for Black Americans focus on personal failure to follow public health advice and lifestyle choices that result in co-morbid conditions (e.g., coronary artery disease and diabetes).^59^ Neither, however, addresses excess exposure, which is structured by institutional racism (and captured in the parameters *β_i→j_*). Indeed, reported mortality rates up to 7 times that of white populations likely reflect considerable underdiagnosis of cases in Black communities,^60^ rather than intrinsic differences in risk of death once infected. In other words, while there is some differential mortality by race for COVID-19 (exacerbated by allostatic load), incidence is likely much higher in Black communities than we appreciate.

### The Symbolic Violence of *R_0_*

Contrary to the way it is often depicted popularly, *R_0_* is not an intrinsic property of a particular pathogen (nor are mortality rates^61^). Rather, the reproduction ratio encapsulates social structure, behavior, and differential risk in a population.^62^ Such risk is often structural,^63^ however, and modeling studies seldom capture oppressive social forces such as institutionalized racism and sexism in their emphasis on ‘objective,’ well-defined parameters. While some scholars attribute this to the inherent conservatism of causal reasoning,^64,65^ it may be more justly described as a form of symbolic violence, referring to the ways naturalized symbols and language sustain relations of oppression.^66–69^ In the case of epidemic modeling, we rarely are presented with racial-justice interventions as ways of preventing and containing outbreaks. Accordingly, this paper has utilized the properties of mathematical models of infectious diseases to illustrate how pandemic containment policy can go beyond the wearing of masks and stay-at-home orders: interventions in risk structure—that is, the way people are enabled or constrained in their associations with others—are crucial to pandemic preparedness, the ability to comply with containment policy once it is decreed, and racial justice in general. Such an amelioration of structural risk can be achieved with reparations.

Since reparations have not been enacted, however, ‘reopening’ American society early (after coronavirus-forced shutdowns) will have a disproportionate adverse mortality effect on Black people, *an effect that is predictable*. Therefore, *de facto*, it resembles a modern Tuskegee experiment, since massive wealth redistribution could avert these deaths, just as penicillin would have in the nearby state of Alabama.^70,71^ As the APM Research Lab has reported, “If they had died of COVID-19 at the same rate as White Americans, about 13,000 Black Americans, 1,300 Latino Americans and 300 Asian Americans would still be alive”^60^—and this is before even 5% of the national population has been infected. The appalling evidence of racism embodied as disproportionate COVID-19 incidence and mortality for Black Americans should add to moral, historical, and legal arguments for reparations for descendants of slaves.

## Data Availability

The authors confirm that the data supporting the findings of this study are available within the article [and/or] its supplementary materials.

